# AMYGDALA VOLUME IS ASSOCIATED WITH ADHD RISK AND SEVERITY BEYOND COMORBIDITIES IN ADOLESCENTS: CLINICAL TESTING OF BRAIN CHART REFERENCE STANDARDS

**DOI:** 10.1101/2023.09.17.23295664

**Authors:** Ádám Nárai, Petra Hermann, Alexandra Rádosi, Pál Vakli, Béla Weiss, János M. Réthelyi, Nóra Bunford, Zoltán Vidnyánszky

**Affiliations:** Brain Imaging Centre, Research Centre for Natural Sciences, Budapest, Hungary; Doctoral School of Biology and Sportbiology, Institute of Biology, Faculty of Sciences, University of Pécs, Pécs, Hungary; Clinical and Developmental Neuropsychology Research Group, Institute of Cognitive Neuroscience and Psychology, Research Centre for Natural Sciences, Budapest, Hungary; Department of Psychiatry and Psychotherapy, Faculty of Medicine, Semmelweis University, Budapest, Hungary

**Author notes:** shared corresponding and senior authors (Nóra Bunford, Zoltán Vidnyánszky).

**Keywords:** brain chart, normative modeling, ADHD, adolescent, MRI

## Abstract

*Background.* Understanding atypicalities in ADHD brain correlates is a step towards better understanding ADHD etiology. Efforts to map atypicalities at the level of brain structure have been hindered by the absence of normative reference standards. Recent publication of brain charts allows for assessment of individual variation relative to age- and sex-adjusted reference standards and thus estimation not only of case-control differences but also of intraindividual prediction. *Methods.* Aim was to examine, whether brain charts can be applied in a sample of adolescents (*N*=140, 38% female) to determine whether atypical brain subcortical and total volumes are associated with ADHD at-risk status and severity of parent-rated symptoms, accounting for self-rated anxiety and depression, and parent-rated oppositional defiant disorder (ODD) as well as motion. *Results.* Smaller bilateral amygdala volume was associated with ADHD at-risk status, beyond effects of comorbidities and motion, and smaller bilateral amygdala volume was associated with inattention and hyperactivity/impulsivity, beyond effects of comorbidities except for ODD symptoms, and motion. *Conclusions.* Individual differences in amygdala volume meaningfully add to estimating ADHD risk and severity. Conceptually, amygdalar involvement is consistent with behavioral and functional imaging data on atypical reinforcement sensitivity as a marker of ADHD-related risk. Methodologically, results show that brain chart reference standards can be applied to address clinically informative, focused and specific questions.

## INTRODUCTION

Attention-deficit/hyperactivity disorder (ADHD), characterized by developmentally-inappropriate inattention, hyperactivity and/or impulsivity as well as functional impairment (1), is an early-onset yet often lifelong, prevalent, and costly disorder. ADHD is typically diagnosed in the early school years (1) and its symptoms persist, in 50% of cases, at a diagnostic (2), and in 65% of cases, at an impairing (3) level into adulthood. Worldwide, ADHD is diagnosed in ∼5-9% of children and adolescents – hundreds of millions of youth (4–6) and ADHD-associated annual costs amount to hundreds of billions of dollars (7).

Attenuating the personal and societal burden of ADHD necessitates improvement of early identification of at-risk individuals and of personalized prevention, which in turn necessitates improvements in understanding ADHD etiology.

Alterations or atypicalities in brain function and structure are at the center of conceptual frameworks explaining ADHD etiology (8–13) and ample data indicate differences between children and adults with and without ADHD in brain volume. Specifically, meta-analytic findings pooling region of interest brain volume (14) and voxel-based morphometry (VBM) (15–18) studies consistently indicate reduced volumes in individuals with ADHD in certain parts of the basal ganglia (caudate, putamen, globus pallidus). Albeit less consistently, results also suggest reduced volumes in the cerebrum, corpus callosum (splenium), insula, medial prefrontal cortex, ventromedial orbitofrontal cortex, and anterior cingulate cortex (14–18). More recently, larger samples allowed for the detection of case-control effect sizes observed in other psychiatric disorders and data showed reduced volumes in ADHD in regions beyond those observed previously, including in the accumbens, amygdala, hippocampus and intracranial volume (ICV) (19).

Indicative of clinical utility, altered brain volume, in turn, is associated with ADHD clinical features in clinical and in the general populations. Greater severity of symptoms is associated in clinical samples with smaller volumes of the caudate, cerebellum, and frontal and temporal gray matter (20) and in general population samples with smaller total volume (21). Both severity of symptoms and the ADHD syndrome are also associated in community samples with slower cortical thinning predominantly in prefrontal cortical regions, bilaterally in the middle frontal/premotor gyri, extending down the medial prefrontal wall to the anterior cingulate; the orbitofrontal cortex; and the right inferior frontal gyrus (22).

Yet, the majority of available data on structural brain differences in children and adults with ADHD have been obtained in case-control designs. Case-control designs, although informative about the extent to which - *at the group level -* individuals with ADHD differ from individuals without ADHD, are less informative about the extent to which *any given individual with ADHD* is atypical. An emerging but robust body of work indicates the need to at least augment if not shift focus from between-groups comparisons to the assessment of differences at the individual level. Specifically, data show considerable within-group heterogeneity both in ADHD and in the general population (23–25) and thus suggest that personalization of behavioral medicine necessitates estimation and knowledge of differences at the individual level.

In case of anthropometric traits such as head circumference, height and weight, individual differences can be quantified against reference standards (i.e., normative growth charts). As disorders like ADHD, autism spectrum disorder (ASD), or schizophrenia are caused/ characterized by atypical brain development, the ability to quantify individual differences is at least comparably if not especially relevant in case of clinical neuroimaging. Yet, until most recently (26,27), no brain development reference standards were available. Capitalizing on advances across neuroimaging and statistical techniques and on availability of large datasets, brain charts have been generated to define reference standards for structural brain measures including global features such as total cortical and subcortical gray matter volume, total white matter volume, and total ventricular cerebrospinal fluid (26) as well as specific estimates for 188 different brain regions (27) across ages and sexes (26,27). Data obtained for the development and validation of brain charts indicate the charts are appropriate for dissecting biological heterogeneity in clinical disorders (e.g., anxiety and depression; ADHD and ASD; and degenerative disorders) both in terms of global features (26) and in summary metrics (representing deviation patterns across individual brain regions) (27) of brain development.

### Current study

As a next step towards demonstrating clinical potential of brain charts, the aim in the current study was to examine whether brain development reference standards can be applied for addressing specific questions in estimating clinical effects in a deeply phenotyped sample. Specifically, aim was to examine whether brain charts can be applied in a sample of adolescents to determine whether atypical brain region volumes predict ADHD at-risk status and severity of inattention (IA) and hyperactivity/impulsivity (H/I), accounting for the effects of symptoms of common comorbidities including anxiety, depression, and oppositional defiant disorder (ODD) as well as the effects of motion.

In addition to estimating effects of total cranial volume, given current conceptual understanding of ADHD etiology (8), findings of earlier meta-analyses (14–19), and findings of recent normative modeling analyses (28), we also modeled effects of individual subcortical regions. Historically, leading conceptual frameworks of ADHD have centered on the prefrontal cortex and its interconnections with the striatum and other subcortical structures in explaining ADHD pathophysiology (8). However, empirical findings only partially support these frameworks; as indicated by the developmental remission of symptomatology, the functional and neural development of the prefrontal cortex parallels ADHD recovery rather than development or onset. ADHD appears more to be a consequence of noncortical dysfunction that does not parallel ADHD remission but manifests early in ontogeny and remains stable into adulthood (8,29).

Psychiatric comorbidity in ADHD is less the exception and more the rule: common comorbidities in ADHD observed across clinical and community samples include anxiety disorders, depression, and ODD, with comorbidity rates around 25% for anxiety (30) 12-50% for depression (31) and 30-50% for ODD (31). To avoid introduction of confounds, comorbidities were accounted for in analyses.

As ADHD diagnostic criteria and corresponding interview and scale measures indicate ADHD is associated with locomotor hyperactivity (though behavioral data are mixed (12)), ADHD may be associated with greater motion in the MRI (32,33) and this may confound measures of brain anatomy (34,35). Thus, effects of motion were also accounted for in analyses.

Analyses were replicated, as in (36), using the output of traditional MRI preprocessing methods (hereafter: raw data) to compare findings obtained using normative modeling with those obtained not applying reference standards.

## METHODS AND MATERIALS

### General Procedure

Data analyzed in the current study were obtained during the first three assessment sessions of the second (baseline, i.e., T1) year of a larger longitudinal project, the (WITHHELD) study.

Exclusionary criteria were cognitive ability ≤the percentile rank corresponding to a full-scale IQ score of 80 (37,38); meeting diagnostic criteria for autism spectrum (severity≥2), bipolar, obsessive-compulsive, or psychotic disorder on the Structured Clinical Interview for DSM-5 Disorders, Clinical Version (SCID-5 CV) (39); neurological illness; and visual impairment (uncorrected, impaired vision <50 cm).

Parents and participants provided written informed consent (and assent) and then participants underwent a series of tests, including assessment of cognitive ability and a clinical interview, followed by genetic sampling and questionnaires (first assessment session) and an EEG measurement and questionnaires (second assessment session), and an MRI measurement (third assessment session). Parents completed questionnaires using the Psytoolkit platform (40,41) and the Qualtrics software, Version June 2020–March 2021 (Qualtrics, Provo, UT). This research was approved by the National Institute of Pharmacy and Nutrition (OGYÉI/27030/2020) and has been performed in accordance with the ethical standards laid down in the 1964 Declaration of Helsinki and its later amendments.

ADHD classification was determined using parent-report on the ADHD Rating Scale-5 (ARS-5) (42). To be classified as at-risk for ADHD, adolescents had to meet a total of ≥4 of the Diagnostic and Statistical Manual of Mental Disorders (5th ed.; DSM-5) ADHD symptoms (from either the IA or the H/I domain).

### Participants

Participants were adolescents from the larger community sample of adolescents oversampled for ADHD who participated in the MRI measurement. The analysis sample for this study was comprised of *N*=140 adolescents.

### Measures

#### Clinical Measures

Anxiety and depression symptoms were assessed by self-report on the Youth Self-Report (YSR) (43), ODD symptoms by parent-report on the Disruptive Behavior Disorders-Rating Scale (DBD-RS) (44), and ADHD risk and severity by parent-report on the ADHD Rating Scale-5 (ARS-5) (42). Prior findings indicate all employed measures have adequate psychometric properties and in the current sample, all exhibited acceptable internal consistency (see Supplemental Information for details).

#### MRI Data Acquisition and Preprocessing

Structural imaging was performed using a magnetization-prepared rapid gradient-echo (MPRAGE) scan with 2-fold in-plane GRAPPA acceleration on a Siemens Magnetom Prisma 3T MRI scanner with the standard Siemens 32-channel head coil using the following parameters: isotropic 1 mm^3^ spatial resolution, repetition time (TR)=2300 ms, echo time (TE)=3 ms, inversion time (TI)=900 ms, flip angle (FA)=9°, field of view (FOV)=2561Z256 mm. Subcortical segmentation of T1-weighted structural images and estimation of morphometric statistics was performed using FreeSurfer 7.1.1 with the built-in probabilistic atlas (45). A novel normative modeling framework (27) was used to calculate deviation *z*-scores for each subcortical volume, total subcortical gray matter volume and left- and right hemispheric mean cortical thickness. The deviation *z*-scores were averaged between hemispheres for the subcortical ROIs. For estimation we used the lifespan_57K_82sites model (27) and adapted it to our own site using an in-house structural MRI dataset of 379 participants (230 female, mean age: 41.95, SD: 19.02, range: 18-80 years). Each scan in the adaptation dataset was performed on the same scanner, using the same MPRAGE protocol as for the analyzed dataset.

### Analytic Plan

Logistic and linear regression models with Elastic-Net regularization (both L1 and L2 penalty terms added) were used to determine whether and how atypical brain morphometry is associated with ADHD at-risk status and IA and H/I severity. Hyperparameter tuning was performed using 5-fold cross validation with 20 l1_ratio values between 0 and 1 on an inverse logarithmic scale and 20 automatically selected values for regularization strength. For the logistic regression classifier, stratified cross validation and balanced class weighting were used. The best hyperparameter combination was selected using Area Under the Receiver Operating Characteristic Curve (AUC) scores for the logistic and coefficient of determination (*R*^2^) scores for the linear regression models. Models were refitted and evaluated on the whole dataset to calculate final coefficients and goodness of fit measures (AUC and *R*^2^ scores). Statistical evaluation of the results was performed using two-tailed permutation tests with 1000 random permutations and an alpha level of .05.

Modeling was performed on both deviation z-scores of normative brain morphometry and on raw morphometric estimates. Since deviation z-scores are inherently corrected for age and sex, age and sex were included as additional covariates when using raw morphometric estimates. Global models included the left- and right hemispheric mean cortical thickness and the total volume of subcortical gray matter. Subcortical models included the volumes of 8 subcortical ROIs (Thalamus, Caudate, Putamen, Pallidum, Hippocampus, Amygdala, Accumbens, Ventral Diencephalon) averaged between hemispheres. All models included the intracranial volume (ICV) as control for the variance in total brain size. To control for the possible effects of participant motion, we included the Euler number (calculated from the FreeSurfer output) as covariate in all models, as it was shown to be highly correlated with manual ratings of motion artifacts (46). Furthermore, based on findings of a recent study (47) performed on our T1-weighted motion controlled dataset (48), SNR derived image quality metrics (IQMs) of MRIQC 0.16.1 (49), namely SNR and SNRd for gray matter (GM), white matter (WM) and cerebrospinal fluid (CSF) were also included as motion covariates in all models. Anxiety, depression, and ODD scores were also controlled for. As ODD was highly correlated with dependent variables and thus could mask effects of other predictors, each model was also fitted without the ODD variable. All features were standardized to aid interpretability of coefficients and modeling was performed in Python (3.11.4) using the scikit-learn (1.3.0) (50) package.

## RESULTS

### Descriptive results

In the analysis sample of *N*=140 adolescents (*M*_age_=15.686, *SD*=1.029; 38% female), families represented a slightly above-average socioeconomic background (average family net income was in the 500 001-700 000 HUF/month range, with average family net income in Hungary being 147 000 HUF/month) (51). For additional descriptive results, see Table 1.

**Table 1.**
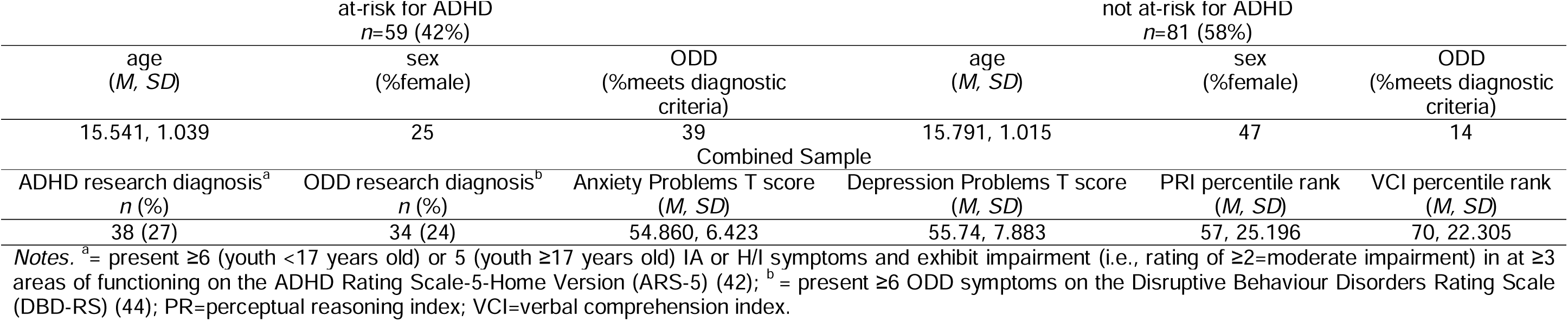
Descriptive results for the analysis sample.

| at-risk for ADHD<br>*n*=59 (42%) |  |  | not at-risk for ADHD<br>*n*=81 (58%) |  |  |
| --- | --- | --- | --- | --- | --- |
| age<br>(*M*, *SD*) | sex<br>(%female) | ODD<br>(%meets diagnostic<br>criteria) | age<br>(*M*, *SD*) | sex<br>(%female) | ODD<br>(%meets diagnostic<br>criteria) |
| 15.541, 1.039 | 25 | 39 | 15.791, 1.015 | 47 | 14 |
| Combined Sample |  |  |  |  |  |
| ADHD research diagnosis<sup>a</sup><br>*n* (%) | ODD research diagnosis<sup>b</sup><br>*n* (%) | Anxiety Problems T score<br>(*M*, *SD*) | Depression Problems T score<br>(*M*, *SD*) | PRi percentile rank<br>(*M*, *SD*) | VCI percentile rank<br>(*M*, *SD*) |
| 38 (27) | 34 (24) | 54.860, 6.423 | 55.74, 7.883 | 57, 25.196 | 70, 22.305 |

*Notes.* <sup>a</sup> = present ≥6 (youth <17 years old) or 5 (youth ≥17 years old) IA or H/I symptoms and exhibit impairment (i.e., rating of ≥2=moderate impairment) in at ≥3 areas of functioning on the ADHD Rating Scale-5-Home Version (ARS-5) (42); <sup>b</sup> = present ≥6 ODD symptoms on the Disruptive Behaviour Disorders Rating Scale (DBD-RS) (44); PR=perceptual reasoning index; VCI=verbal comprehension index.

Of adolescents at-risk for ADHD, *n*=12 were currently prescribed ADHD pharmacological treatment. Of these, 7 were prescribed stimulants, 6 were prescribed nonstimulants (1 was prescribed both), and two took a 24-hour medication hiatus prior to the MRI session, three did not, and seven did not indicate whether they took a hiatus.

Independent samples Mann-Whitney U tests indicated at-risk and not at-risk groups differed on IA, H/I, and ODD scores (all *p*_FDRcorr_<.001) with those at-risk scoring higher on these measures. Groups did not differ on age, SES, perceptual or verbal reasoning, anxiety, or depression scores (all *p*_FDRcorr_>.084).

### Normative modeling results

We initially used global measures of brain morphometry to investigate whether cortical thickness or subcortical gray matter volume are associated with ADHD at-risk status. Global models included the normative volume of total subcortical gray matter, the normative mean cortical thickness (separately for the two hemispheres), total ICV, motion covariates, anxiety and depression scores. The model including ODD scores predicted ADHD at-risk status (AUC=.879, *p*=.001), with a negative association of total subcortical gray matter volume (coef=-.742, *p*=.009) and a positive association of ODD scores (coef=1.63, *p*=.001) with ADHD at-risk status. The effect of non-interest of CSF SNR was also significant (coef=-.806, *p*=.001). Despite not reaching a significant model fit (AUC=.71, *p*=.057), the model without ODD scores predicted ADHD at-risk status, with a negative association of total subcortical gray matter volume (coef=-.306, *p*=.013) with ADHD at-risk status. The effects of non-interest of CSF SNR (coef=-.364, *p*=.011) and GM SNRd (coef=-.149, *p*=.039) were also significant. Cortical thickness was not associated with ADHD at-risk status in either model (all *p*s>.215).

To further investigate the effect of subcortical gray matter volume on ADHD at-risk status, we defined subcortical models with the normative volume of subcortical ROIs as predictors, along with ICV, motion covariates, anxiety, and depression scores. In estimating ADHD at-risk status, the model including ODD scores predicted ADHD at-risk status (AUC=.858, *p*=.001), with a negative association of bilateral amygdala volume (coef=-.136, *p*=.017), a negative association of bilateral ventral diencephalon (coef=-.069, *p*=.047) and a positive association of ODD scores (coef=.836, *p*=.001) with ADHD at-risk status (Fig. 1). The effect of non-interest of CSF SNR was also significant (coef=-.209, *p*=.009). Despite not reaching a significant model fit (AUC=.714, *p*=.107), the model without ODD scores predicted ADHD at-risk status, with a negative association of bilateral amygdala volume (coef=-.312, *p*=.007) with ADHD at-risk status. The effect of non-interest of CSF SNR was also significant (coef=-.252, *p*=.011).

**Figure 1.**
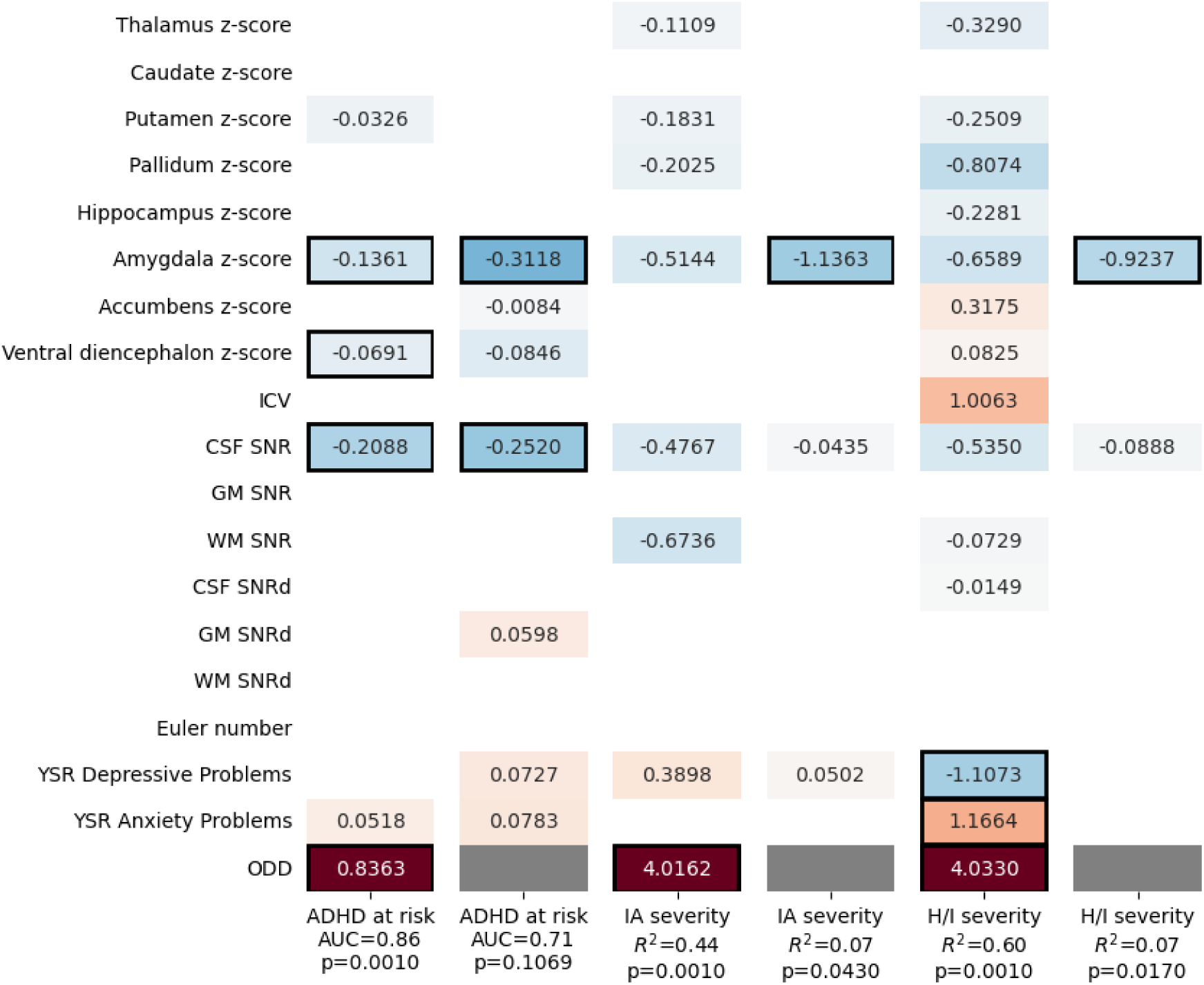
Modeling ADHD at risk status, IA and H/I severity using normative volumes of subcortical ROIs. Cells with black frames indicate significant (*p*<.05) coefficients and white cells indicate zero coefficients. Gray cells mark where the ODD predictor variable was omitted from the model.

In estimating severity of symptoms, models with ODD scores predicted IA (*R*^2^=.442, *p*=.001) and H/I (*R*^2^=.601, *p*=.001), with an association of ODD with IA (coef=4.016, *p*=.001) and H/I (coef=4.033, *p*=.001) and also of anxiety (coef=1.166, p=0.037) and depression (coef=-1.107, *p*=0.033) scores with H/I. The models without ODD also predicted IA (*R*^2^=.069., *p*=.043) and H/I (*R*^2^=.073, *p*=.017), with a negative association of bilateral amygdala volume with IA (coef=-1.136, *p*=.003) and H/I (coef=-.924, *p*=.001).

### Raw modeling results

Subcortical models were recreated using the raw volumes of subcortical ROIs to assess whether and how findings obtained using normative modeling differ from those obtained not applying reference standards. In estimating ADHD at-risk status, the model including ODD scores predicted ADHD at-risk status (AUC=.876, *p*=.001), with a positive association of sex (boys scored higher; coef=.315, *p*=.007) and of ODD scores (coef=.850, *p*=.001) with ADHD at-risk status (Fig. 2). The effects of non-interest of CSF SNR (coef=-.304, *p*=.013) and GM SNRd (coef=.102, *p*=.043) were also significant. The model without ODD scores predicted ADHD at-risk status (AUC=.776, *p*=.005) with a negative association of bilateral amygdala volume (coef=-.384, *p*=.007) and a positive association of sex (boys scored higher; coef=.531, *p*=.003) with ADHD at-risk status. The effects of non-interest of CSF SNR (coef=-.266, *p*=.023) and GM SNRd (coef=.208, *p*=.013) were also significant.

**Figure 2.**
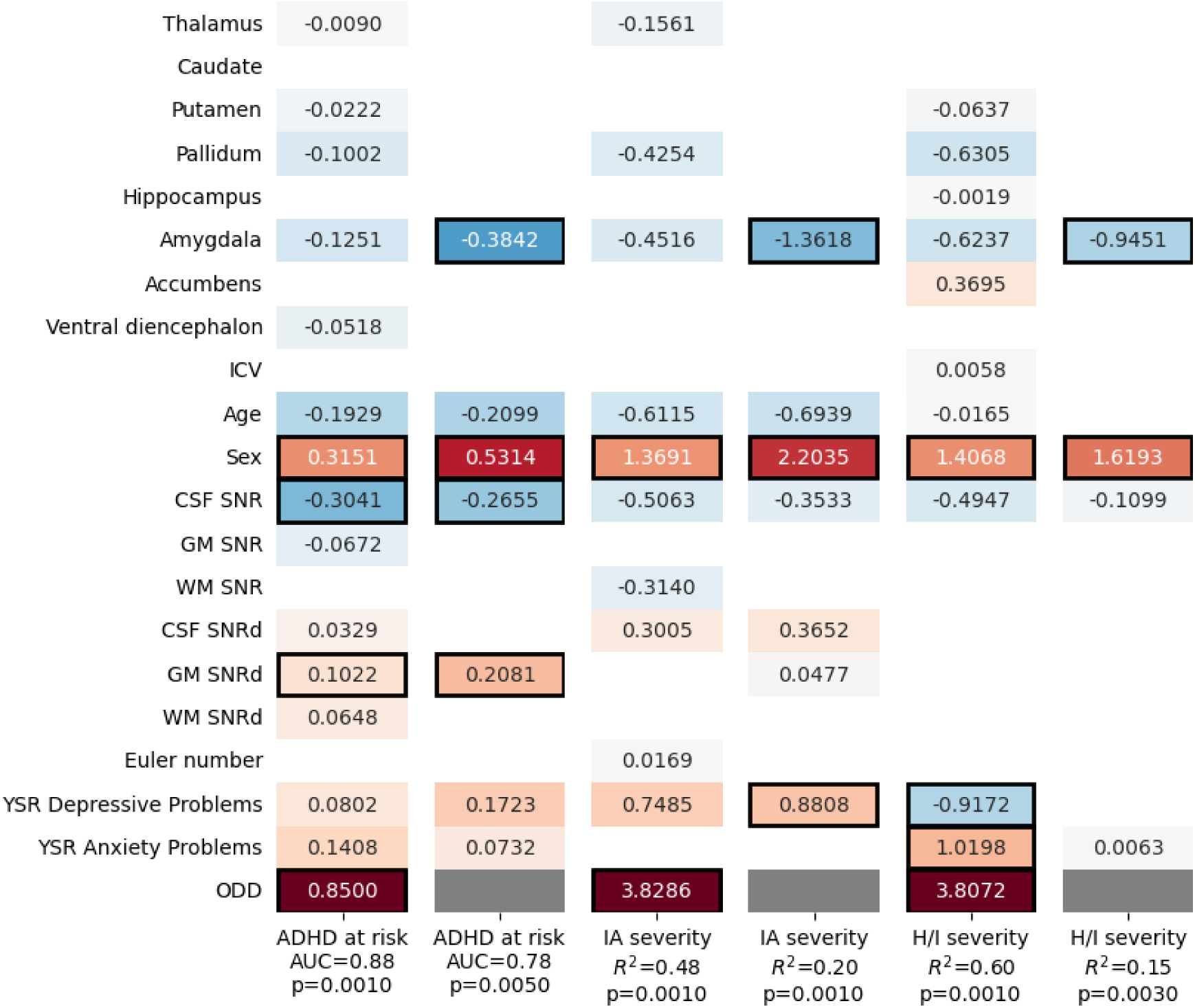
Modeling ADHD at risk status, IA and H/I severity using raw volumes of subcortical ROIs. Cells with black frames indicate significant (*p*<.05) coefficients and white cells indicate zero coefficients. Gray cells mark where the ODD predictor variable was omitted from the model.

In estimating severity of symptoms, models with ODD scores predicted IA (*R*^2^=.478, *p*=.001) and H/I (*R*^2^=.597, *p*=.001), with an association of ODD with IA (coef=3.827, *p*=.001) and H/I (coef=3.807, *p*=.001); of sex with IA (boys scored higher; coef=1.369, *p*=.005) and H/I (coef=1.407, *p*=.013); and of anxiety (coef=1.020, p=0.047) and depression (coef=-.917, *p*=0.037) scores with H/I. The models without ODD predicted IA (*R*^2^=.204., *p*=.001) and H/I (*R*^2^=.148, *p*=.003), with a negative association of bilateral amygdala volume with IA (coef=-1.362, *p*=.001) and H/I (coef=-.945, *p*=.003); a positive association of sex with IA (boys scored higher; coef=2.204, *p*=.001) and H/I (coef=1.619, *p*=.001); and a positive association of depression with IA (coef=.881, *p*=0.021).

## DISCUSSION

Data in the current study indicate that relative to age- and sex-adjusted brain development norms, atypical amygdalar volume is associated with ADHD risk and severity in adolescents. Specifically, above and beyond the effects of the symptoms of common comorbidities including anxiety, depression and ODD as well as motion, reduced bilateral amygdala volume is associated with ADHD at-risk status and, above the effects of the symptoms of anxiety and depression as well as motion, reduced bilateral amygdala volume is associated with inattention and hyperactivity/impulsivity severity. These findings have conceptual and methodological implications.

Findings involving the amygdala are consistent with recent results of a cross-sectional mega-analysis showing smaller amygdala volumes in individuals with relative to without ADHD (19). Conceptually, the current results suggest that ADHD risk is predicted, above and beyond anxiety, depression, and ODD severity, by individual differences in the volume of a brain region implicated in affective processing and reinforcement sensitivity.

The amygdala has been historically associated with fear conditioning and processing of negative and positive emotions (52) and amygdala volume has been implicated in negative affectivity (53). A body of work indicates ADHD is associated with high parent- and teacher report negative affectivity and self-report neuroticism (54–58). Yet, it has been argued that negative affectivity and neuroticism may be general markers of psychopathology (59), and as rating scale measures of negative affectivity include items reflecting anger and irritability, negative affectivity may be more closely related to the externalizing spectrum generally, and ODD specifically, than to ADHD *per se* (60). Indeed, in some samples, compared to the number of children with ADHD characterized by low control or high surgency, only a very small subgroup are characterized by high negative affect (61).

ADHD is also associated with characteristics relevant to positive affectivity (54,62,63) and specifically with positive affectivity (64,65) as well as dysregulation of negative emotions (66,67). Evidence indicates the association between ADHD and positive affectivity is independent of relevant comorbidities (65,68) but, similar to negative affectivity, there is debate about whether dysregulation of negative emotions is more closely related to externalizing and internalizing comorbidities or to ADHD per se (66).

The amygdala is also implicated in reinforcement sensitivity: albeit not a primary region for processing of reward, it plays a central role in coding affective information about environmental stimuli, thereby providing contextual information used for adjusting motivational level (69). Given its critical interactions with the striatum for stimulus-reward associations, it has even been proposed that the amygdala’s role in stimulus-reward learning might be just as important as its role in fear conditioning and processing of negative affect (52). Metabolic PET studies report amygdalar activation in contexts involving potential punishment and rewards (70) and fMRI studies report amygdalar activation to potential reward (71). Of note, controlling for arousal, direct comparison of amygdalar response to punishment vs. reward often reveals no differences, suggesting that amygdalar BOLD signal reflects arousal rather than value (69).

Data across behavioral, neuroimaging, and neurophysiological levels of analysis indicate atypical reinforcement sensitivity is a candidate ADHD intermediate phenotype. Intermediate phenotypes are heritable biological markers located on the etiologic path in-between genetic predisposition and observable symptoms. Given limitations of the mechanistically and phenotypically heterogeneous clinical phenotype (72), intermediate phenotypes have been proposed as adjunct, or supplementary foci for research aimed at determining ADHD etiology. Specifically, relative to the clinical phenotype, by virtue of their homogeneity, intermediate phenotypes are associated with a fewer number of susceptibility traits and a narrower outcome set and thus have better explanatory and predictive power (68,73). In case of reinforcement sensitivity and ADHD, behaviorally, children with ADHD are hypersensitive to reinforcing effects of rewards (74,75), exhibit a stronger preference for immediate compared to delayed rewards (76,77), make more risky decisions to obtain rewards (78), and show steeper temporal discounting (79,80). Functional neuroimaging data show that individuals with, relative to controls without ADHD, exhibit dorsolateral prefrontal (81) and striatal (82) hypoactivation to reward anticipation, but occipital (83) as well as prefrontal and striatal hyperactivation to reward receipt (84–86). Neurophysiological results suggest anticipation of and initial response to reward concurrently (68) and prospectively (65) modulate the association between dispositional positive affectivity and IA and H/I severity and show promise as prospective ADHD biomarkers of substance use indices of prognosis (73). The association between ADHD and atypical reinforcement sensitivity is independent of developmental effects (86) and relevant comorbidities (65,68).

Taken together, it is likely that the herein observed association between amygdala volume and the ADHD clinical phenotype reflect, rather than differences relevant to affective processing, ADHD-linked differences relevant to reinforcement sensitivity. As such, the current findings potentially provide further, albeit indirect support for atypical reinforcement sensitivity as a candidate ADHD intermediate phenotype. That reduced amygdalar volume is associated with ADHD risk and inattention and hyperactivity/impulsivity severity suggests that in addition to atypicalities at the level of behavior and brain function relevant to reinforcement processing – atypicalities at the level of brain structure in a region implicated in reinforcement processing – may also be markers of ADHD (risk).

Of note, that the current findings were observed even when accounting for the effects of comorbidities underscores the utility of biological measures for predicting clinical outcome above and beyond more cost-effective alternatives (e.g., clinical and demographic measures) (87).

Methodologically, these results show that recently published brain chart reference standards can be applied to address, in specific populations, clinically informative, focused and specific questions. Following others (36), we compared findings obtained using normative modeling to those obtained using raw data and essentially, results were replicated. Of note, amygdala effects in presence of ODD were only supported when using normative modeling, underscoring the superiority of this novel approach relative to traditional approaches in identifying biological markers.

### Directions for future research

We examined whether individual differences in brain volume predict ADHD at-risk status and severity. Although there is considerable utility in focusing on subclinical manifestations of the disorder, given emerging evidence that subclinical levels are associated with functional, negative outcomes (88) and may better capture such associations in girls whose ADHD is often more subtle (89), it will be important to determine whether current findings replicate across different classification thresholds, e.g., in predicting ADHD diagnostic status.

We established utility of the brain chart in predicting differences in symptomatology and it will be a next step to assess such utility in detecting additional indices of ADHD clinical functioning, including functional impairment or prognosis.

We did not directly test whether smaller amygdalar volume is related to atypical reinforcement sensitivity as an ADHD intermediate phenotype; this will need to be determined in a within-group design, i.e., whether in individuals with the ADHD clinical phenotype differences in amygdala volume predict differences in reinforcement sensitivity.

In the current study, we applied regularized logistic and linear regression to uncover the effects of subcortical ROI volumes on ADHD risk status. Although predictive modeling of ADHD risk status with an independent test set or using nested cross validation could provide further insight into the underlying relationships in our data and potentially provide a tool for predicting ADHD risk status from new structural MRI scans, our dataset was deemed too small for such analyses. Nevertheless, this could be an important next step for our research.

## Data Availability

All data produced in the present study are available upon reasonable request to the authors

## Author Contribution Statements

**ÁN:** Conceptualization; Methodology; Software; Validation; Formal analysis; Investigation; Data Curation; Writing - Original Draft; Writing - Review & Editing; Visualization

**PH:** Data Curation; Writing - Review & Editing

**AR:** Investigation; Data Curation; Writing - Review & Editing; Project administration

**PV:** Data Curation; Writing - Review & Editing

**BW:** Methodology; Software; Formal analysis; Data Curation; Writing - Review & Editing

**JMR:** Supervision; Writing - Review & Editing

**NB:** Conceptualization; Methodology; Formal analysis; Writing - Original Draft, Writing - Review & Editing, Supervision; Project administration; Funding acquisition

**ZV:** Conceptualization; Methodology; Writing - Review & Editing; Supervision; Project administration; Funding acquisition

## Data Availability

The datasets generated and/or analyzed during the current study are available from the corresponding authors on reasonable request.

