## Supplement for "AMYGDALA VOLUME IS ASSOCIATED WITH ADHD RISK AND SEVERITY BEYOND COMORBIDITIES IN ADOLESCENTS: CLINICAL TESTING OF BRAIN CHART REFERENCE STANDARDS"

**SUPPLEMENTAL INFORMATION**

### *Adolescent Self-Report Measures*

**Anxiety Problems and Depressive Problems.** The Youth Self-Report (YSR) (1) is a 112-item self-report questionnaire for adolescents (ages 11–18) assessing aspects of adaptive and impaired functioning. The YSR measures adaptive functioning through *competence scales*: academic performance, activities, and social competence and impaired functioning via *Diagnostic and Statistical Manual of Mental Disorders (DSM)-oriented scales*: anxiety problems, depressive problems, somatic problems, attention-deficit/ hyperactivity problems, oppositional defiant problems, and conduct problems; as well as *syndrome scales*: anxious/depressed, depressed/withdrawn, somatic complaints, attention problems, social problems, thought problems, aggressive behavior, rule-breaking behavior, externalizing problems and internalizing problems. Respondents rate items on a 3-point scale (0 – 'Not True', 1 – 'Somewhat or Sometimes True', 2 – 'Very True or often True').

Prior findings indicate both the original (1) and the Hungarian translation of the YSR (2) have acceptable psychometric properties.

In the current study, the anxiety and the depressive problems subscales were used in analyses.

**Oppositional Defiant Disorder.** The Disruptive Behavior Disorders-Rating Scale (DBD-RS) (3) is a 45-item parent- and teacher-report measure of the presence and severity of DSM-III-R ADHD symptoms (9 inattentive symptom items and 9 hyperactivity/impulsivity symptom items), oppositional defiant disorder (8 items), and conduct disorder symptoms (15 items). Parents and teachers rate items on a four-point scale ranging 0 (not at all) to 3 (very much), with higher scores indicating more severe symptoms. In the current study, the parent-report form was used and the oppositional defiant disorder items were of interest. Because items reflect DSM-III-R symptom wording, items were modified to match DSM-5 symptom wording (American Psychiatric Association, 2022).

Prior findings indicate both the original (4) and the Hungarian translation of the DBD-RS (2) have acceptable psychometric properties.

In the current sample, the ODD subscale exhibited ω=.916 internal consistency. In the current study, the ODD subscale was used in analyses.

### *Parent-report measures*

**ADHD risk and severity.** The ADHD Rating Scale-5 (ARS-5) (5) is a 30-item parent- and teacher-report measure of the past 6-month presence and severity of DSM-5 ADHD symptoms (9 inattentive symptom items and 9 hyperactivity/impulsivity symptom items) and functional impairment across six domains: relationship with significant others (family members for the home version), relationship with peers, academic functioning, behavioral functioning, homework performance and self-esteem (2×6 impairment items, with one set corresponding to inattention and one to hyperactivity/impulsivity). Parents and teachers rate items on a four-point scale ranging in case of symptoms from 0 (never or rarely) to 4 (very often) and in case of impairment from 0 (no problem) to 3 (severe problem), with higher scores indicating more severe symptoms and impairment. The ARS-5 is comprised of two symptoms scales, Inattention and Hyperactivity-Impulsivity, and a Total Scale. The ARS-5 is suitable for ages 5-17 years, with separate forms for children (5-10 years) and adolescents (11-17 years) and age-appropriate and DSM-5 compatible descriptions of symptoms. In the current study, the adolescent home (i.e., parent-report) version was used.

Prior findings indicate both the original (5) and the Hungarian translation of the ARS-5 (2,6,7) has acceptable psychometric properties.

In the current sample, the ARS-5 total (ω=.950), as well as the inattention (ω=.947) and hyperactivity/impulsivity (ω=.910) subscales exhibited acceptable internal consistency. In the current study, the ARS-5 was used for ADHD classification and the total score as well as the IA and H/I subscales were used in statistical analyses.
